# Therapeutic Exercise in Temporomandibular Disorders: A Scoping Review protocol

**DOI:** 10.1101/2025.01.10.25320171

**Authors:** Gabriele Aimini, Paolo Mastromarchi, Lorenzo Lo Russo, Marco Chiodini, Roberto Giova Bazzacchi, Noemi Corbetta, Federico Amateis, Stefano Salvioli

**Affiliations:** Craniomandibular institute, Milano, Italy; Scuola Universitaria per la Svizzera Italiana, DEASS, Manno, Switzerland; Department of Allied Health Professions, Sheffield Hallam University, Sheffield, UK; IMTA member; Varese, Italy; Magenta, Italy; Department of Maxillo-Facial Surgery, Santi Paolo e Carlo Hospital, University of Milan, 20122 Milan, Italy; Department of Neurosciences, Rehabilitation, Ophthalmology, Genetics, Maternal and Child Health, University of Genoa, Campus of Savona, Italy; Department of Neuroscience, Rehabilitation, Ophthalmology, Genetics and Maternal and Child Sciences (DINOGMI), University of Genoa, Campus of Savona, Italy

**Keywords:** temporomandibular joint dysfunction, mandibular disease, physical exercise, active movement, exercise training

## Abstract

Temporomandibular Disorders (TMDs) are musculoskeletal dysfunctions affecting the temporomandibular joint and masticatory muscles, causing pain and functional limitations. The etiology of TMDs is multifactorial, including biological factors, parafunctional habits (such as bruxism), and psychological conditions. This study aims to map and synthesize existing evidence on therapeutic exercise for TMDs.

**Inclusion criteria:** All primary studies,randomized controlled trials (RCTs), prospective and retrospective observational studies, case studies will be considered.

**Methods:** This scoping review will be performed in accordance with the Joanna Briggs Institute (JBI) methodology. MEDLINE, Cochrane Central, Scopus, CINAHL Complete, Embase, PEDro and will be searched from inception until now. Additional records will be identified through searching in Google Scholar(the first 5 pages were examined) and Open Grey,, clinicaltrials.gov, Trialsearch.who.int and the reference lists of all relevant studies. No study design, publication type, or data restrictions will be applied. Two reviewers will independently screen all abstracts and full-text studies for inclusion. A data collection form will be developed by the research team to extract the characteristics of the studies included. A tabular and accompanying narrative summary of the information will be provided.

**Conclusion:** This will be the first scoping review to provide a comprehensive overview and the results will add significant information for clinicians in the management of TMDs with therapeutic exercise. In addition, any gaps in knowledge on the topic will be identified. The results of this research will be published in a peer-reviewed journal and will be presented at relevant (inter)national scientific events.

## Introduction

Temporomandibular disorders (TMDs) are a series of musculoskeletal dysfunctions affecting the temporomandibular joint or the masticatory muscles and/or related structures (1,2).

TMDs can be characterized by pain in the mandibular, oral, facial, and periauricular areas, limitations of passive or active mouth opening, and joint sounds (3,4); pain can also spread to several neighbouring structures and generate other symptoms such as tinnitus, otalgia, headache, and cervicalgia (5,6,7).

TMDs are the main cause of pain of the orofacial and head region, after those of dental and periodontal origin (8). A recent study by Valesan (2020) reported that TMDs affect 5-12% of the adult population and 11% of children and adolescents, TMDs were considered the most common cause of chronic pain of non-dental origin in the orofacial area. In both age groups, the most frequent dysfunction was disc dislocation, followed by degenerative joint pathology (9).

Furthermore, TMDs are more frequent in the female population, with women having twice the risk of developing them than men. Interestingly, this prevalence in the female gender in a ratio of 2:1 can be found in all diagnostic groups identified according to the research diagnostic criteria for temporomandibular disorders (RDC/TMDs) Axis I, more specifically 2.09:1 in the muscle disorders group, 1.6:1 in the disc displacement group and 2.08:1 in the arthralgia/arthritis/arthrosis group (10).

There seems to be a direct correlation between the most severe cases of TMDs and a lower quality of life in terms of altered taste, discomfort during eating, voice changes, chronic pain resulting in absence from work, loss of energy, insomnia, restriction of physical activity, emotional disturbances, anxiety, stress, depression, reduced economic income due to extended need for medical services, and predictable social repercussions (11).

The etiology of TMDs is multifactorial varying from physical predisposition factors to para- functional habits such as cheek, lip or tooth biting (bruxism), from comorbidity with other neck or head disorders to psychological factors (12, 14–17).

Due to the complexity and heterogeneity of clinical presentations, there is no unique method in treating these patients, the literature suggests multimodal physiotherapy treatments combining therapeutic exercise, education, manual therapy, cognitive behavioural therapies, laser therapy, and dry needling (4,18,20,21).

Idáñez-Robles et al. (2023) in a recent systematic review with meta-analysis investigated the variability of treatment proposals for patients with TMDs evaluating the available evidence on the effectiveness of physiotherapy in improving pain and passive and active opening of the mouth (22).

Also, Arribas-Pascual et al. addressed this topic with an umbrella and systematic mapping review with meta-analysis. They concluded that manual therapy, therapeutic exercise, and low-level laser therapy effectively reduced pain intensity and improved maximal mouth opening in TMDs (23).

Despite the existence of substantial evidence supporting the efficacy of physiotherapy interventions for TMDs, there is an absence of guidelines delineating the dosage and type of exercise to be administered in daily clinical settings. Instead, there is considerable heterogeneity in the intervention content, encompassing a wide spectrum of approaches, from general exercises involving the entire body, grounded in postural correction, to targeted exercises for the temporomandibular joint, masticatory muscles, or the cervical spine.

Therefore, this scoping review aims to map the available studies on therapeutic exercise for TMDs in terms of the type of exercise proposed and its characteristics: intensity, dosage, frequency, and mode of administration.

### Research objective (s)

1. A compendium of therapeutic exercises for TMDs will be compiled. The compendium will encompass active movements, resistance control exercises, stretching, posture and muscle strengthening exercises, and other types of active exercises.
2. The characteristics of the proposed exercises for TMDs will be investigated, including the mode of administration, intensity, dosage, facilitation, posology, direction of movement, progressions, and outcome measures.
3. The study will examine the types and characteristics of exercise most frequently used within TMDs subgroups, including active mouth opening movements, lateral movements, protrusion movements, and motor control exercises.
4. The secondary objective is to map by year of publication, study design, geographical distribution of TMDs, classification and terminology used to define TMDs.

### Inclusion criteria

Studies will be eligible for inclusion if they meet the Population, Concept, and Context (PCC) criteria:

#### Population

studies with adult participants (from 18 years), diagnosed by any type of health professional (e.g. doctor, dentist, physiotherapist…), with any type of TMDs.

#### Concept

All types of exercise (e.g., exercise with or without supervision, active exercise, passive-active exercise, postural exercise for TMDs, jaw exercise for TMD, neck exercise for TMDs, shoulder exercise for TMDs, holistic approach for TMD).

#### Context

All contexts in which exercise is administered, such as 1:1 delivery, group, outpatient, home, self-administered, delivered by any clinician.

#### Sources

all primary studies, randomized controlled trials (RCTs), prospective and retrospective observational studies, and case studies will be considered.

#### Language

English, Italian, French, Spanish.

### Exclusion criteria

#### Population

Studies of persons with fractures, genetic abnormalities, post-operative conditions, infections, and tumour conditions affecting the TMJ will be excluded, well as studies in asymptomatic subjects.

Studies that do not meet the specific inclusion PCC criteria will be excluded. Systematic reviews, conference abstracts, narrative reviews and non-peer-reviewed studies will be excluded.

## Methods

The proposed scoping review will be conducted following the JBI methodology for scoping reviews (24). It will be reported according to the PRISMA-ScR (25).

### Search strategy

An initial limited search of MEDLINE will be undertaken to identify articles on the topic and then index terms used to describe the articles will be used to develop a full search strategy for MEDLINE (see Table 1, which displays the search strategy for MEDLINE database). The search strategy, including all identified keywords and index terms, will be adapted for use in Cochrane Central, Scopus, CINAHL Complete, Embase, Pedro.

In addition, Google Scholar (the first 5 pages were examined), Open Grey, clinicaltrials.gov, trialsearch.who.int the reference lists of all relevant studies will be searched.

### Study selection

Once the search strategy is completed, search results will be collated and imported to online the software Rayyan (rayyan.qcri.org) (26). Duplicates will be removed using Rayyan. The review process will consist of two levels of screening: (1) a title and abstract screening and (2) a full-text screening. All labels for the studies excluded through the criteria defined by the PCC will be saved on Rayyan itself. For both levels, two authors (GA, and LLR) will independently screen the articles, and a third author (FA) will resolve any emerging conflicts.

The reasons for the exclusion of any full-text paper will be recorded and reported in a scoping review appendix. The results of the search will be reported in full in the final scoping review and presented with a Preferred Reporting Items for Systematic Reviews and Meta-analyses (PRISMA) flow diagram (27).

### Data extraction

Key data and information from selected articles will be collected including authors, country, year of publication, type of publication, type of TMDs, outcome measures, type of exercise and posology.

This form will be reviewed by the research team and pre-tested by all reviewers before implementation to ensure that the form can most accurately accommodate all the data that will be extracted. The presentation of results is usually an iterative process during scoping reviews; additional data may be added to this module based on subgroups that may emerge from the analysis of the included studies (28,29).

### Data synthesis

The results of the research will be presented in three ways:

1. The process will use numerical framing to select relevant studies, with a detailed and mapped description to ensure reproducibility. The extracted data will be summarised in tables and graphs. Outcome mapping will follow an iterative approach, with additional categories added as necessary (29).
2. The framing of the themes will be used to produce a thematic synthesis. This will be related to the key concepts and themes relevant to the research questions and according to the subgroups (e.g. different TMD exercises, dosage, type of treatment) and others that may emerge.
3. The use of visual framing, which includes the design of contemporary and comprehensible graphics will be used to facilitate the synthesis of the extracted results, thereby enabling the reader to engage in critical analysis.

## Data Availability

All data produced in the present work are contained in the manuscript
All data produced in the present study are available upon reasonable request to the authors

## Acknowledgements

Not applicable.

## List of abbreviations

JBI: Joanna Briggs Institute;
PCC: Population-concept context;
TMDs: Temporo-mandibular disorders;
PRISMA: Preferred Reporting Items for Systematic Reviews and Meta-analyses;
PRISMA-ScR: The Preferred Reporting Items for Systematic reviews and Metanalyses extension for Scoping Reviews;
RCTs: randomized controlled trials,
RDC/TMDs: research diagnostic criteria for temporomandibular disorders

## Appendix 1

((((((((((Temporomandibular Joint[MeSH Terms]) OR (Temporomandibular Joint[Text Word])) OR (Temporomandibular Joint Dysfunction Syndrome[MeSH Terms])) OR (Temporomandibular Joint Dysfunction Syndrome[Text Word])) OR (Craniomandibular Disorders[MeSH Terms])) OR (Craniomandibular Disorders[Text Word])) OR (Mandibular Diseases[MeSH Terms])) OR (Mandibular Diseases[Text Word])) OR (Jaw Diseases[MeSH Terms])) OR (Jaw Diseases[Text Word])) AND (((((((((((((((((Exercise[MeSH Terms]) OR (Exercise[Text Word])) OR (Motor Activity[MeSH Terms])) OR (Motor Activity[Text Word])) OR (Physical Exercise[MeSH Terms])) OR (Physical Exercise[Text Word])) OR (Exercise Training[MeSH Terms])) OR (Exercise Training[Text Word])) OR (Muscle Stretching Exercises[MeSH Terms])) OR (Muscle Stretching Exercises[Text Word])) OR (active movement[MeSH Terms])) OR (active movement[Text Word])) OR (joint mobilization[Text Word])) OR (Rehabilitation[MeSH Terms])) OR (Rehabilitation[Text Word])) OR (Physical Therapy Modalities[MeSH Terms])) OR (Physical Therapy Modalities[Text Word]))

